# Value-Based Integrated Care: A Systematic Literature Review

**DOI:** 10.1101/2023.10.10.23296735

**Authors:** Evelien S. van Hoorn, Lizhen Ye, Nikki van Leeuwen, Hein Raat, Hester F. Lingsma

## Abstract

**Background:** Healthcare services worldwide are transforming themselves into value-based organizations. Integrated care is an important aspect of value-based healthcare (VBHC), but practical evidence-based recommendations for the successful implementation of integrated care within a VBHC context are lacking. This systematic review aims to identify how value-based integrated care (VBIC) is defined in literature, and to summarize the literature regarding the effects of VBIC, and the facilitators and barriers for its implementation.

**Methods:** Embase, Medline ALL, Web of Science Core Collection and Cochrane Central Register of Controlled Trails databases were searched from inception until January 2022. Empirical studies that implemented and evaluated an integrated care intervention within a VBHC context were included. Non-empirical studies were included if they described either a definition of VBIC or facilitators and barriers for its implementation. The Rainbow Model of Integrated Care (RMIC) was used to analyse the VBIC interventions. The quality of the articles was assessed using the Mixed Methods Appraisal Tool.

**Results:** After screening 1328 titles/abstract and 485 full-text articles, 24 articles were included. No articles were excluded based on quality. One article provided a definition of VBIC. Eleven studies reported -mostly positive-effects of VBIC, on clinical outcomes, patient-reported outcomes, and healthcare utilization. Nineteen studies reported facilitators and barriers for the implementation of VBIC; factors related to reimbursement and IT infrastructure were reported most frequently.

**Conclusion:** The concept of VBIC is not well defined. The effect of VBIC seems promising, but the exact interpretation of effect evaluations is challenged by the precedence of multicomponent interventions, multiple testing and generalizability issues. For successful implementation of VBIC, it is imperative that healthcare organizations consider investing in adequate IT infrastructure and new reimbursement models.

## 1. Background

Nowadays, integrated care is often seen as the future direction for the development of healthcare systems driven by the aging population, increase in patients with comorbidity and the associated increase in healthcare expenditure. (1, 2). Integrated care can take many different forms, and there is no unifying definition (1, 3–6). The definition of integrated care is dependent on the different views and expectations of the various stakeholders (7, 8). What unifies the different definitions, however, is that integrated care is an approach to overcome care fragmentation leading to improved patient outcomes and experiences with care(8, 9).

To successfully implement integrated care, it is essential to understand its complexity. Different taxonomies have been developed to guide healthcare professionals, managers, policymakers, researchers and other stakeholders to differentiate and analyse the different forms of integrated care (7, 8, 10). Those taxonomies typically describe the type of integration (i.e. professional, organizational), the level at which integration occurs (i.e. macro-, meso-, micro-), the degree of integration (i.e. from informal linkages to more managed care coordination and fully integrated teams or organizations), the process of integration (i.e. how integrated care is organized and managed) and the breadth of integration (i.e. to a whole population or specific group) (8).

In order to provide more sustainable healthcare, healthcare services worldwide are transforming themselves into value-based organizations (11, 12). By implementing value-based healthcare (VBHC), healthcare organizations aim to maximize value for patients by achieving the best patient outcomes at the lowest possible costs (13, 14). Integrated care is an important aspect within the VBHC framework; integrated practice units (IPUs) and the integration of care delivery across multiple separate facilities are two of the core pillars of VBHC (13). In an IPU, care is delivered by a dedicated, multidisciplinary team who takes responsibility for the full cycle of care for a specific condition, encompassing outpatient, inpatient and rehabilitative care, as well as supporting services (13). Members of an IPU see themselves as one organizational unit and share a common administrative and scheduling structure. An essential element of integrated care within the VBHC framework, described in theory, is that IPUs routinely measure outcomes, cost, care processes and patient experience using a common platform and accept joint accountability for the results (13, 15).

In the current literature, several reviews have been performed to provide healthcare organizations with practical and evidence-based recommendations for the successful implementation of integrated care. Reviews have summarized the literature on how integrated care is implemented(1), the facilitators and barriers for its implementation (16, 17), and its effectiveness (2, 18, 19). Until now, no overview of the literature exists to identify those elements for integrated care within a VBHC context.

This systematic review aims to provide practical evidence-based recommendations for the successful implementation of integrated care within a VBHC context. To achieve this, we aim to identify how integrated care within a VBHC context, in other words value-based integrated care (VBIC), is defined in the current literature. Furthermore, we aim to summarize the results of evaluations of the effects of VBIC, and to summarize the literature regarding the facilitators and barriers of its implementation.

## 2. Methods

### 2.1 Search strategy

This review was conducted in line with the Preferred Reporting Items for Systematic Reviews and Meta-Analysis (PRISMA) guidelines (20). The electronic databases Embase, Medline ALL, Web of Science Core Collection and Cochrane Central Register of Controlled Trials were systematically searched for relevant articles from the date of inception of each database until January 15^th^ 2022. To identify publications that reported on VBIC, the literature search included search terms related to both VBHC and integrated care. Since there is no unambiguous definition for integrated care, synonyms such as comprehensive care, coordinated care and multidisciplinary care were included within the search. Synonyms and other terms related to VBHC were also incorporated within the search. The search terms were adequately adjusted for each database and included both registered and non-registered index terms. Further details of the search strategy are available in the Supplementary materials. The protocol was registered in the PROSPERO database (registration number CRD42021259025).

### 2.2 Eligibility criteria and article selection

To be eligible for this review, publications had to meet the following criteria: 1) description of an empirical study, 2) covering a healthcare context, 3) written in English or Dutch, 4) description of VBHC or integrated care (including any spelling variation and synonyms) in the introduction or method section, and 5) provide a definition for VBIC, describe the effects of VBIC or mention facilitators and barriers for its implementation. Theoretical articles (e.g. commentaries) and articles without an available full text (e.g. conference abstracts) were excluded. One exception was made to the eligibility criteria. Non-empirical studies were included if they provided a definition of VBIC or mentioned facilitators and barriers for its implementation to ensure all relevant publications were included within this review.

All articles were screened against the eligibility criteria in two phases; first, the titles and abstracts were screened, followed by the full-text screening. Both the title and abstract, and full-text screening was performed by two independent reviewers. When there were conflicts about whether an article met the inclusion criteria, a third reviewer was consulted for a third opinion and discrepancies were discussed until consensus was reached. The article screening was performed using *Covidence*(21).

### 2.3 Quality assessment

The methodological quality of the included articles was appraised independently by two reviewers using the Mixed Methods Appraisal Tool (MMAT). The MMAT permits the appraisal of five different study designs; 1) qualitative research, 2) randomized controlled trials, 3) non-randomized studies, 4) quantitative descriptive studies, and 5) mixed methods studies (22). This allowed the use of one appraisal tool for all included studies within this systematic review. The MMAT consist of two screening questions and five questions per study design. All questions can be answered with “Yes”, “No”, or “Cannot tell”. Responding “No” or “Cannot tell” on the screening questions indicates that the study is not an empirical study and further appraisal may not be feasible or appropriate (22). This review, therefore, only assessed the methodological quality of the empirical studies. Any discrepancies in the quality assessment were resolved by consulting the third reviewer.

### 2.4 Data extraction and analysis

All included studies were analysed qualitatively and data were extracted on the following items: definition of VBIC, the study and intervention characteristics, all outcome measures and results of the VBIC intervention, and the facilitators and barriers for its implementation. The VBIC interventions were categorized using the Rainbow Model of Integrated Care (RMIC)(23). The RMIC distinguishes six integration dimensions (clinical, professional, organizational, system, functional and normative integration) (Table 1). The first four dimensions describe the type of integration and level at which integrated care can occur: macro-(system) level, meso-(organizational and professional) level and micro-(clinical) level. The other dimensions, functional and normative integration, describe the mechanisms, or in other words facilitators, that support the implementation of integrated care. (23, 24)

**Table 1:** Integrated care dimensions of the Rainbow Model of Integrated Care.

| Level | Dimension | Description |
| --- | --- | --- |
| Micro | Clinical integration | The coordination of person-focused care in a single process across time, place and discipline |
| Meso | Professional integration | Inter-professional partnerships based on shared competences, roles, responsibilities and accountability to deliver a comprehensive continuum of care to a defined population |
| Meso | Organizational integration | Inter-organizational relationships (e.g., contracting, strategic alliances, knowledge networks, mergers), including common governance mechanisms, to deliver comprehensive services to a defined population |
| Macro | System integration | A horizontal and vertical integrated system, based on a coherent set of (informal and formal) rules and policies between care providers and external stakeholder for the benefit of people and populations |
| Micro, meso, macro | Functional integration | Key support functions and activities (i.e. financial, management and information systems) structured around the primary process of service delivery to coordinate and support accountability and decision-making between organizations and professionals in order to add overall value to the system. |
| Micro, meso, macro | Normative integration | The development and maintenance of a common frame of reference (i.e. shared mission, vision, values and culture) between individuals, professional groups and organizations. |

Adopted from Valentijn et al. (23, 24)

## 3. Results

### 3.1 Search results

After deduplication, the combined search yielded 1328 unique articles. After the title and abstract screening phase, 485 records were screened on full text. After full text inspection 461 articles were excluded for the following reasons: the article was not in English or Dutch (n=2), no full text was available (n=27), the article did not mention VBHC and integrated care in the introduction or method section (n=156), or the article was not empirical and/or did not describe a definition, facilitators and barriers or effects of VBIC (n=276). At the end, 24 articles met the inclusion criteria (Figure 1).

**Figure 1:**
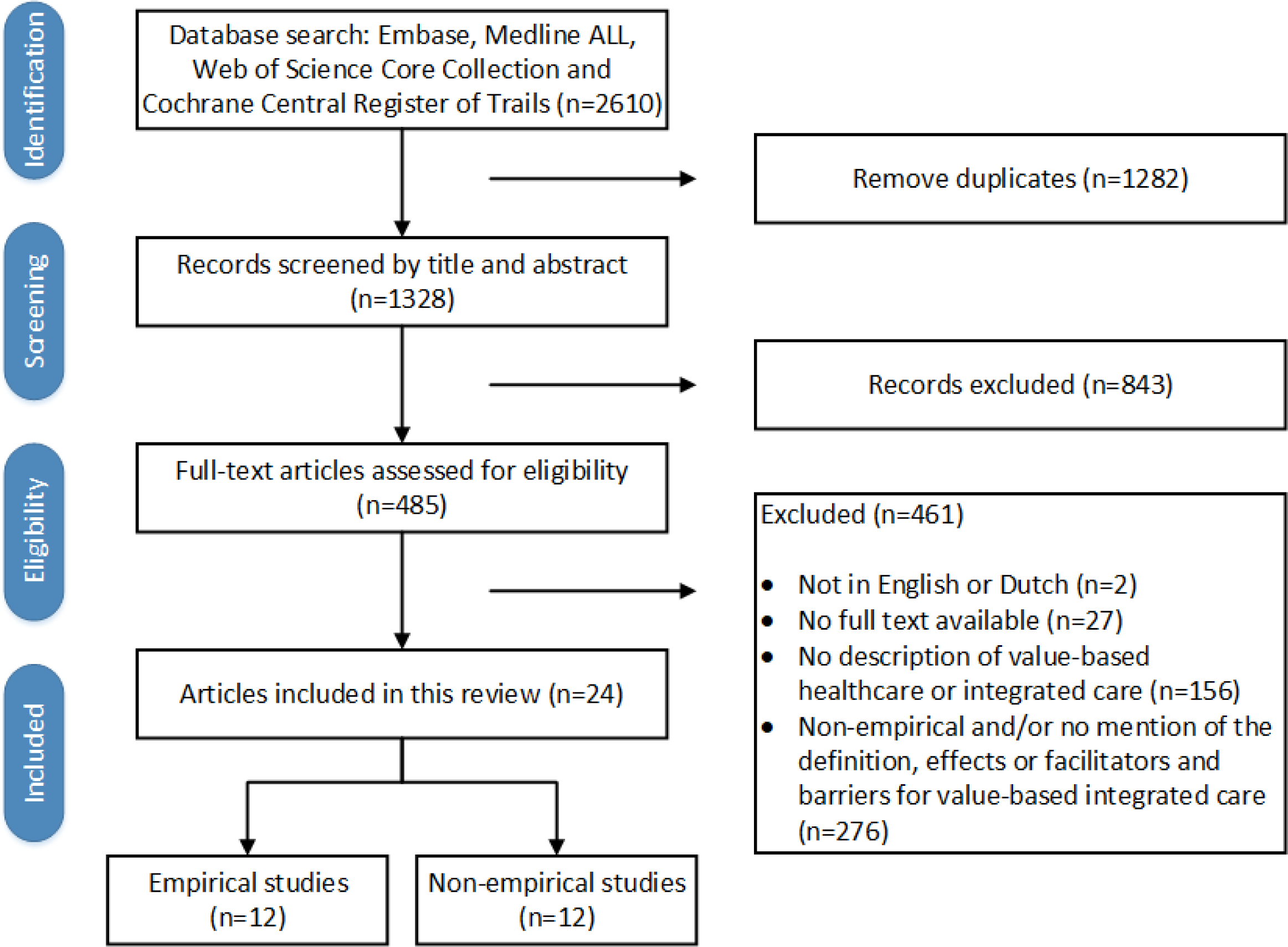
Flowchart depicting the article selection.

### 3.2 Study characteristics and quality

The included articles were published between 2013 and 2021. Seventy-one percent (n=17) of all included studies were conducted in the United States, 8% (n=2) in the Netherlands and the remaining 21% (n=5) in Italy, Spain, Sweden, the United Kingdom, and Taiwan. Fifty percent (n=12) of the included publications described an empirical study, of which almost all performed a quantitative analysis (n=11). Quality assessment was performed for all empirical studies. Of these studies, seven were categorized as non-randomized studies (25–31), three as randomized controlled trails (32–34), one as qualitative study (35) and one as a quantitative descriptive study (36). Almost all articles had a good methodological quality. One article(36) had a questionable methodological quality; almost all questions of the MMAT were answered with “No” or “Cannot tell”. This article was not excluded since it contained relevant information about the facilitators and barriers for VBIC.

### 3.3 The definition of value-based integrated care

Within the included articles, one study provided an explicit definition of VBIC. Valentijn et al. defined value-based integrated care as “patients’ achieved outcomes and experience of care in combination with the amount of money spent by providing accessible, comprehensive and coordinated services to a target population” (37 p.2). Two other articles referred to this definition(38, 39). All other articles did not specify or mention the term VBIC. Those articles used a combination of integrated care synonyms and VBHC to describe VBIC. The most commonly mentioned integrated care synonyms were IPU’s (26, 29, 35, 36, 39–44), multidisciplinary teams (25, 38, 45, 46), multidisciplinary or interdisciplinary care (28, 31, 33), team-based care (30, 47) and working together across disciplines or institutions (27, 48).

### 3.4 Interventions and effects of value-based integrated care

#### 3.4.1 Value-based integrated care interventions

Twelve articles described the implementation and evaluation of an integrated care intervention within a VBHC context (Table 2). The VBIC interventions consisted of multiple components, targeted different patient populations and occurred in different settings. The interventions were implemented in primary care(30, 31, 33, 34), primary and secondary care(36), secondary care(27–29, 32) or tertiary care(25, 26, 35). According to the Rainbow Model of Integrated Care, the VBIC interventions can be classified as clinical(30, 31, 33, 34), professional(26, 28, 29, 32, 35, 36), organization(27) and system integration(25).

**Table 2:**
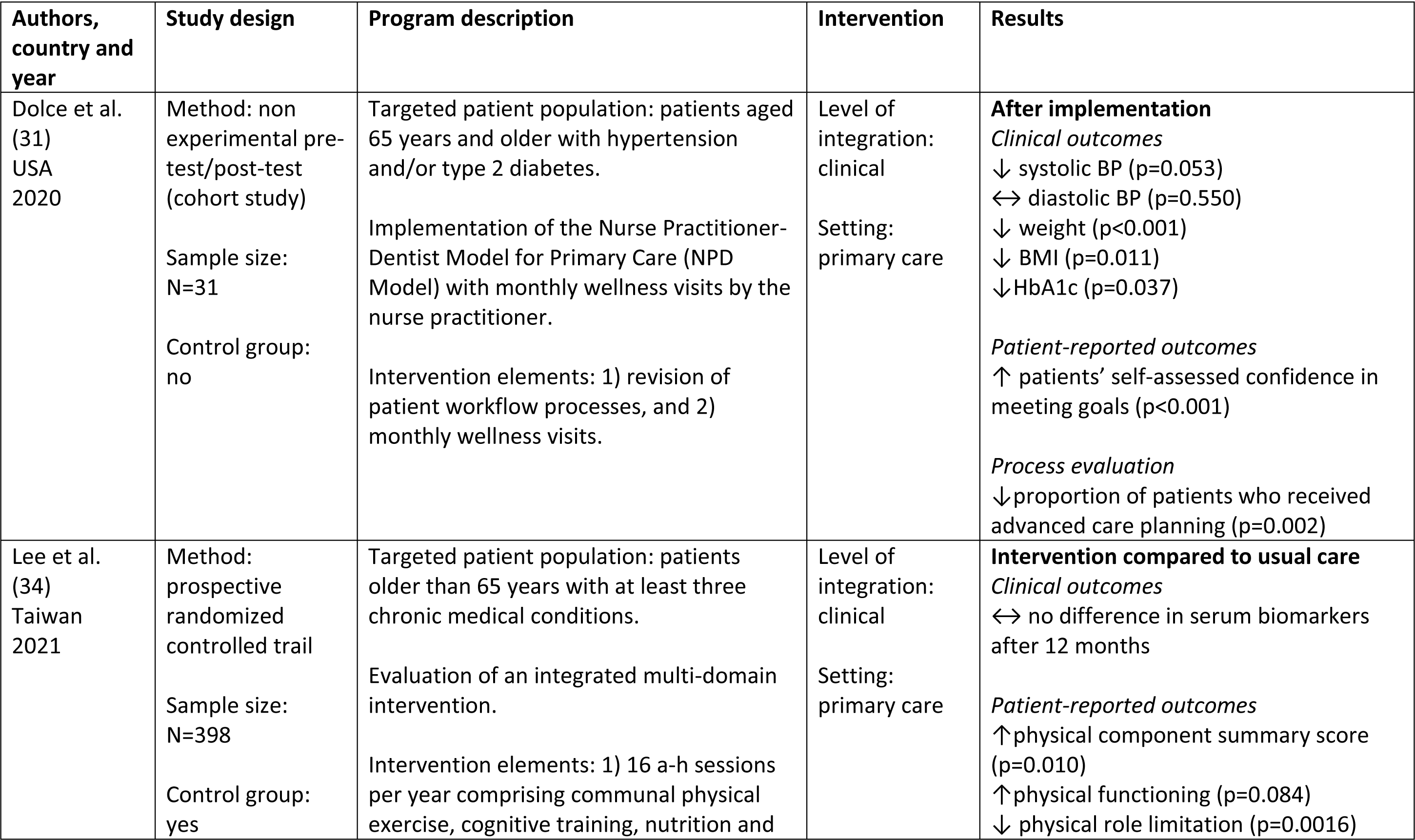
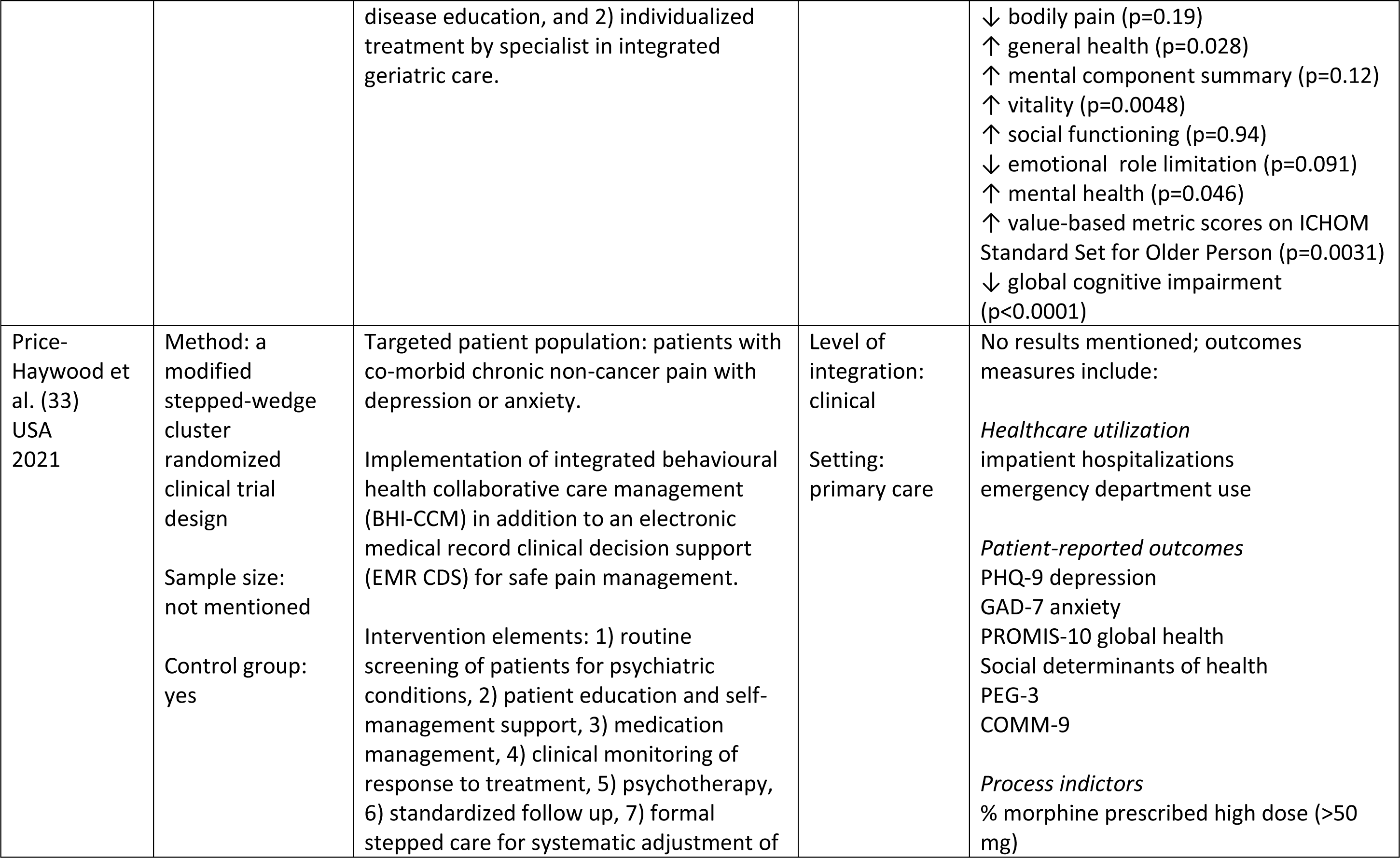
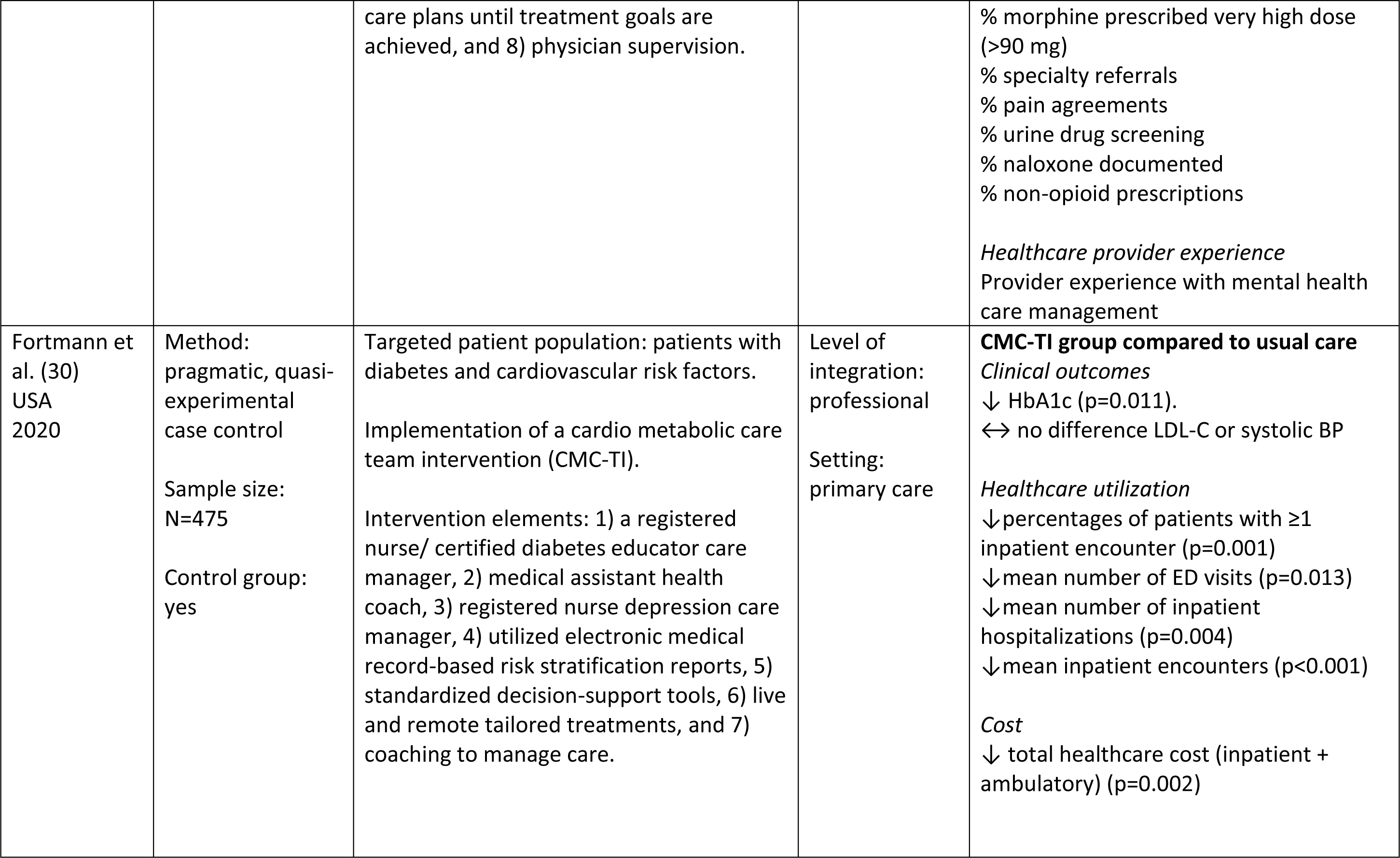
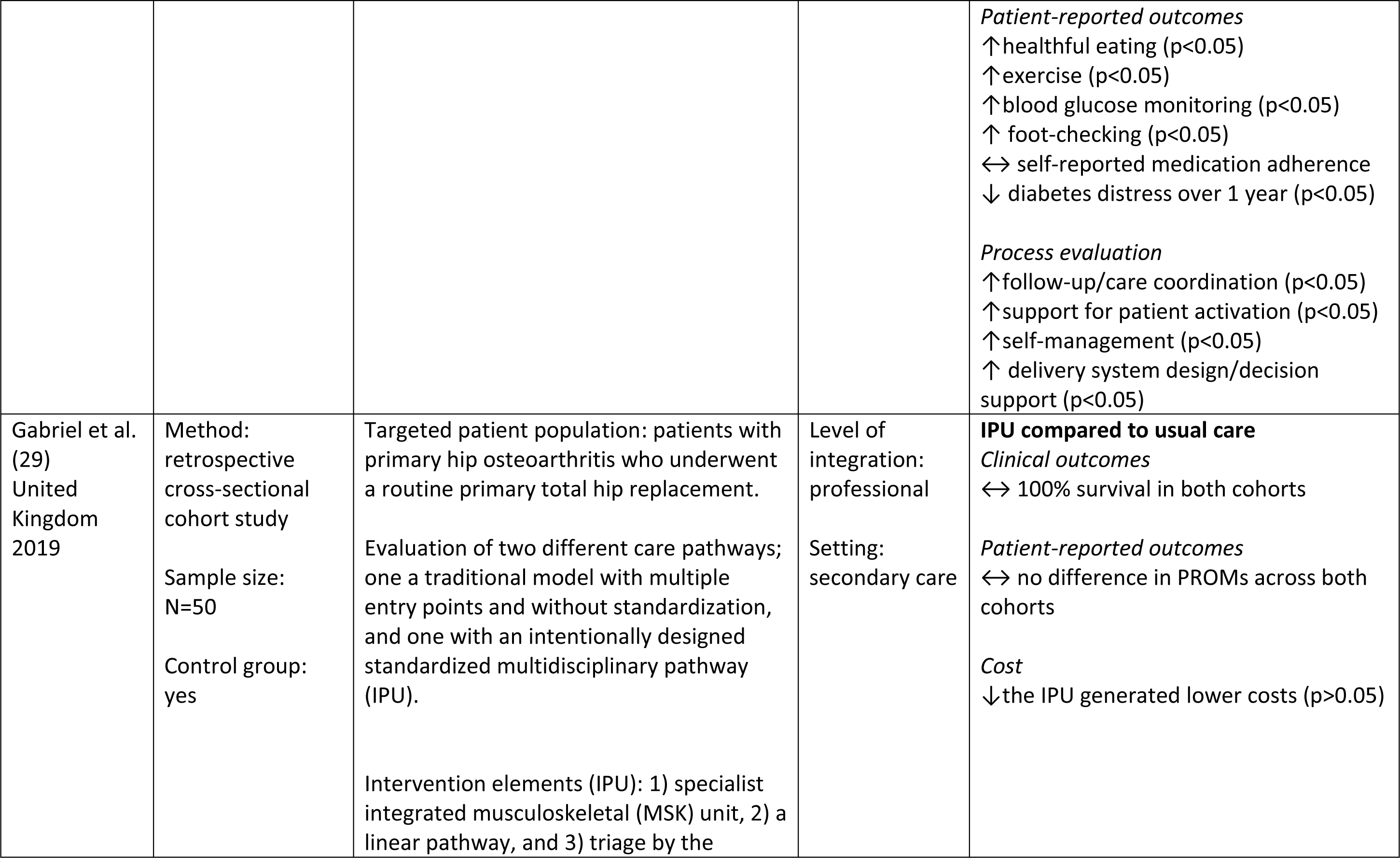
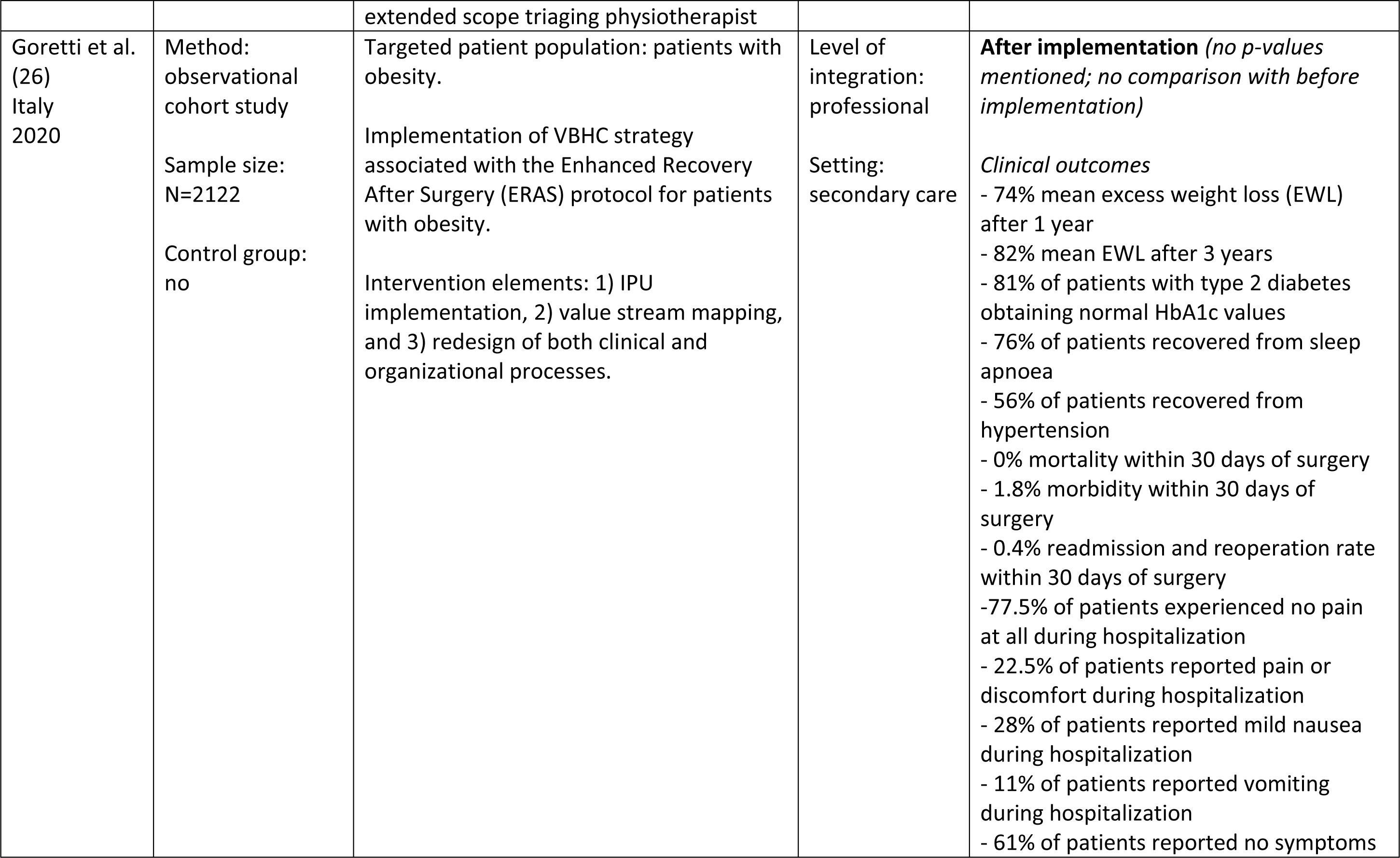
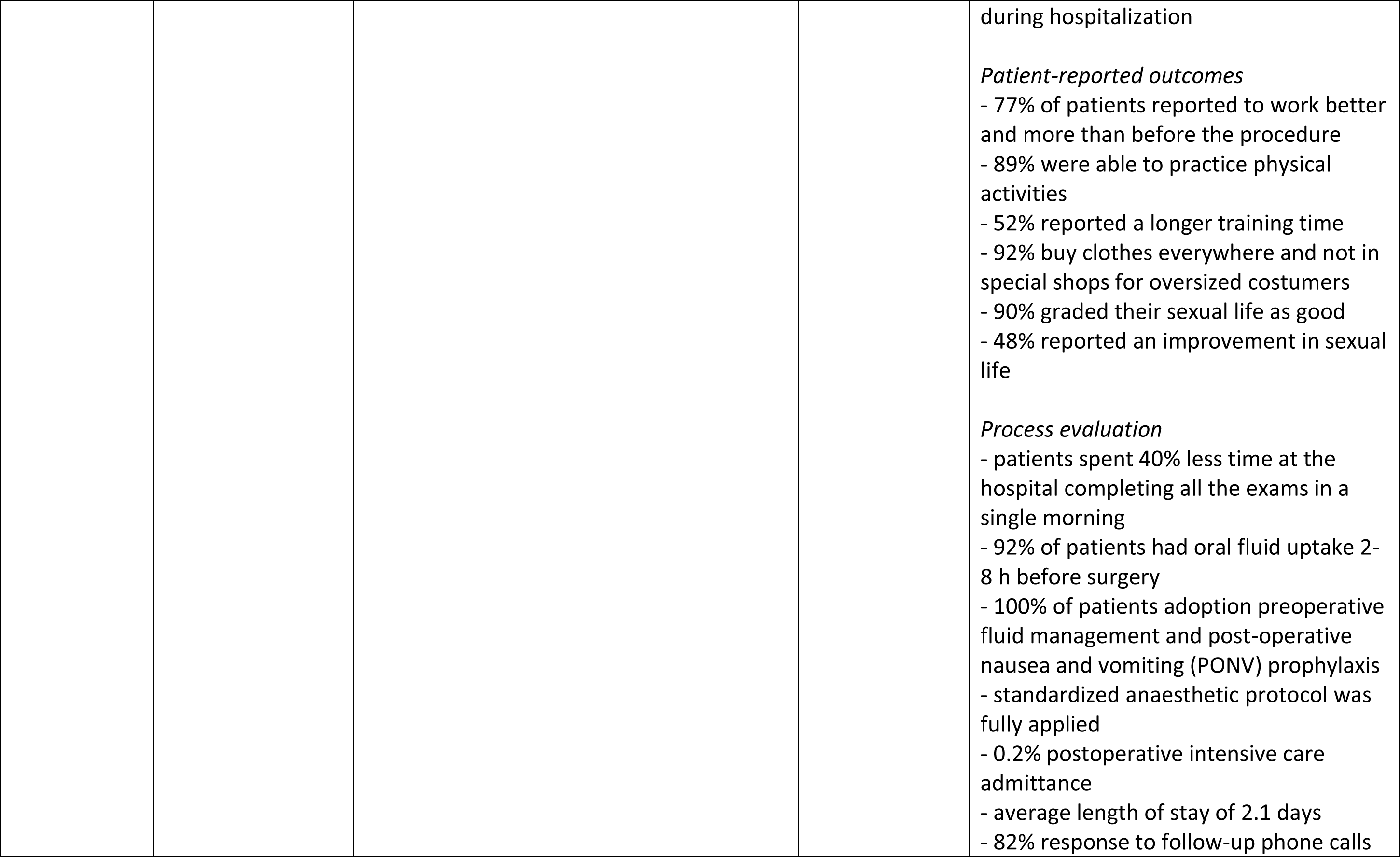
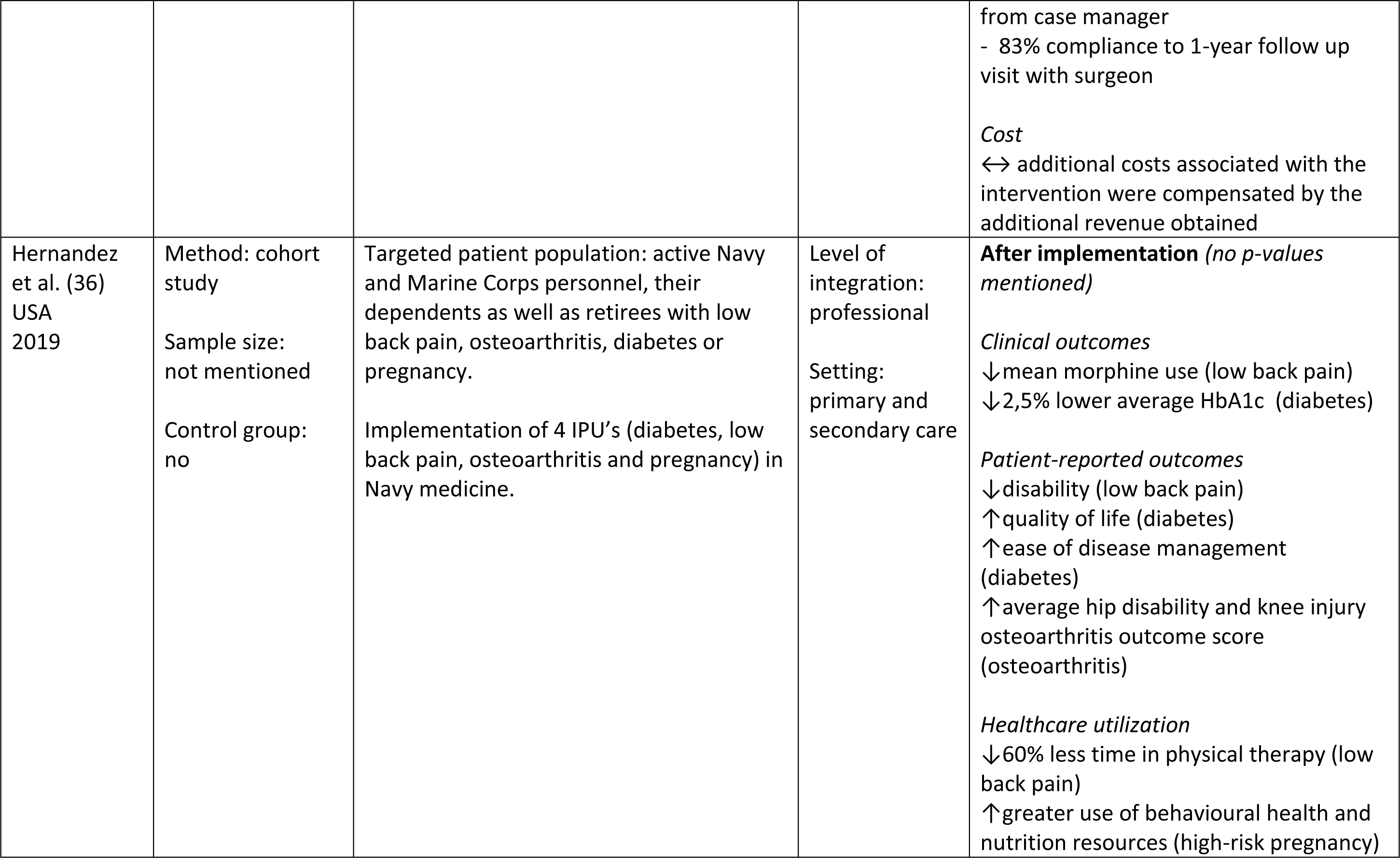
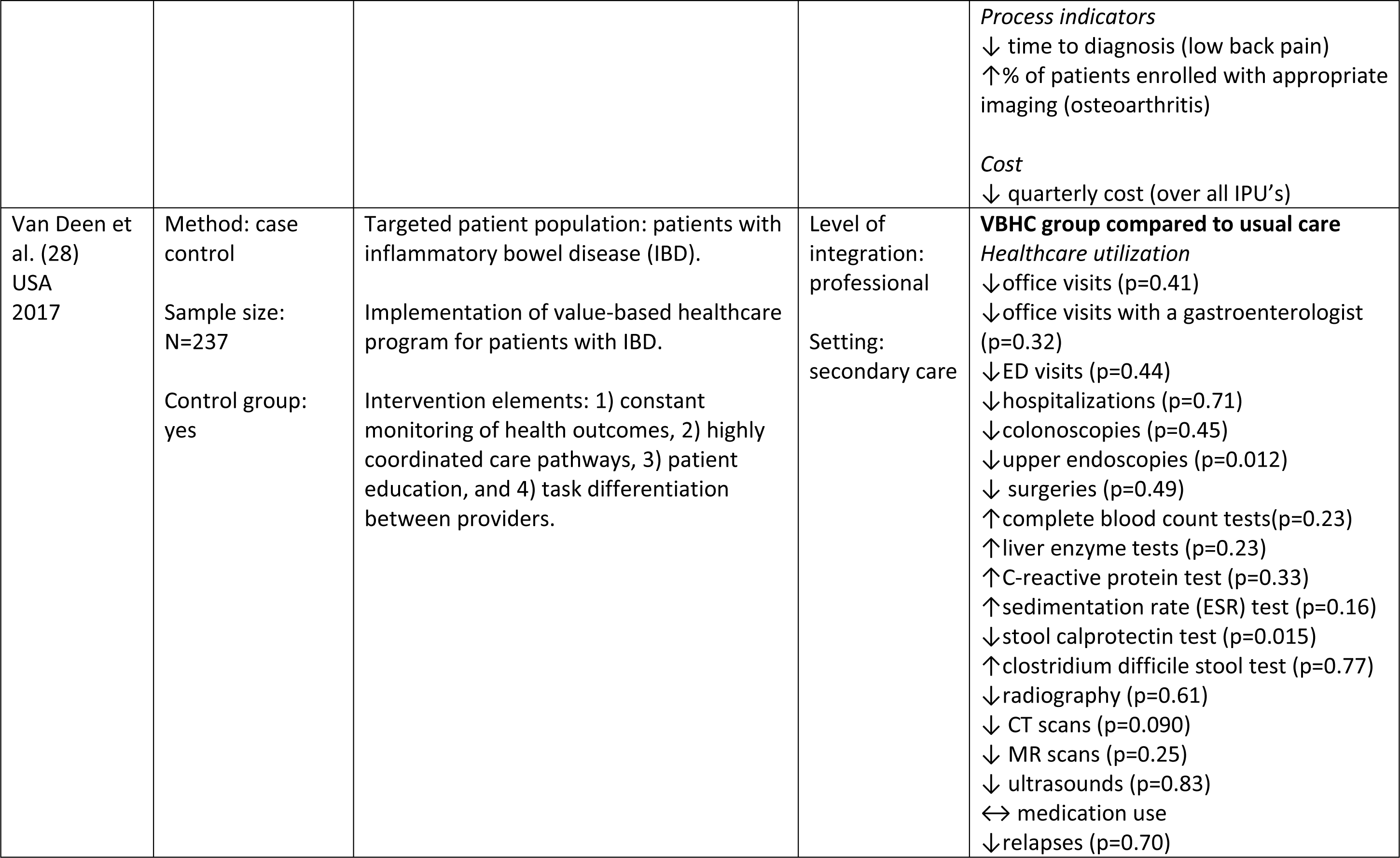
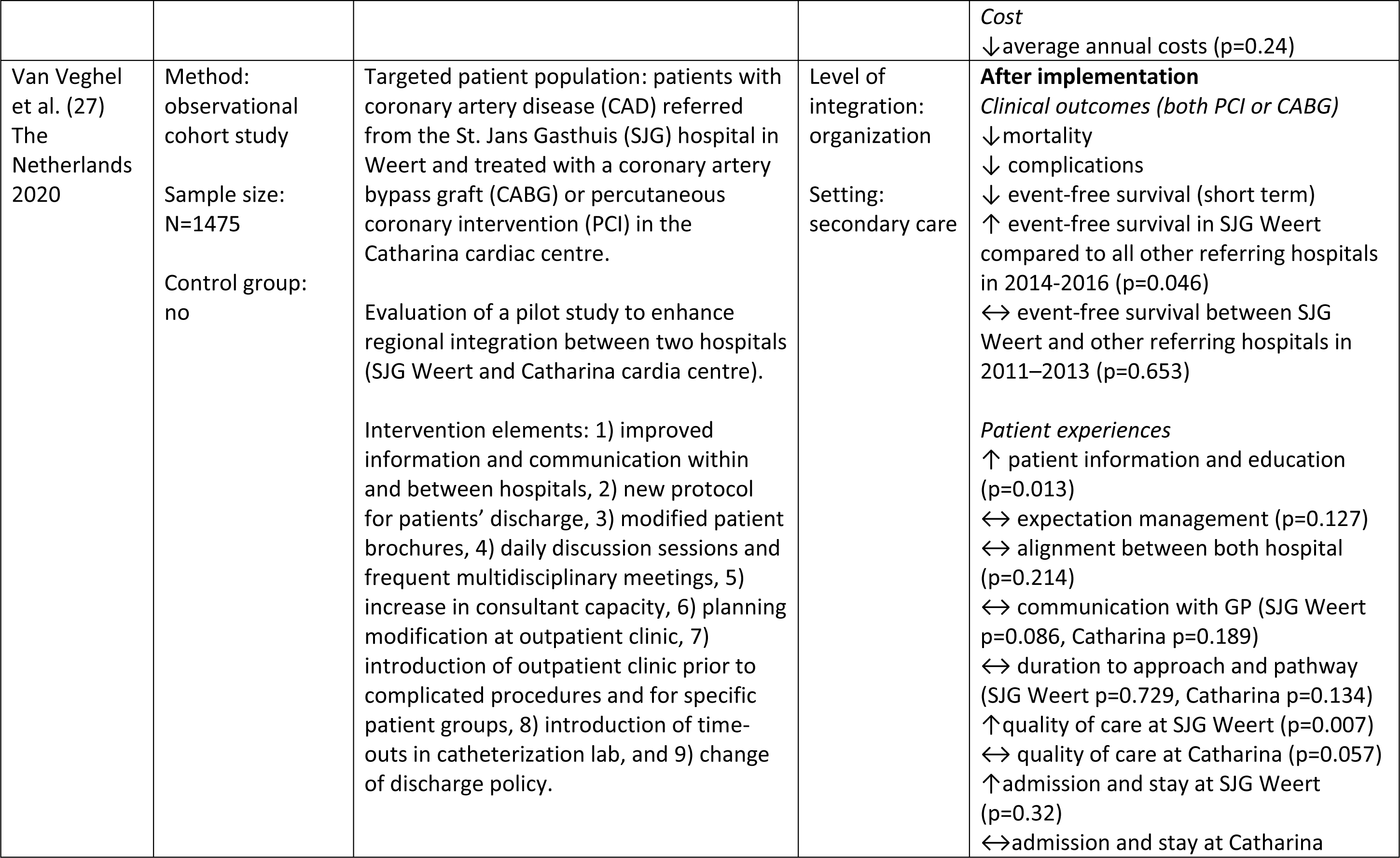
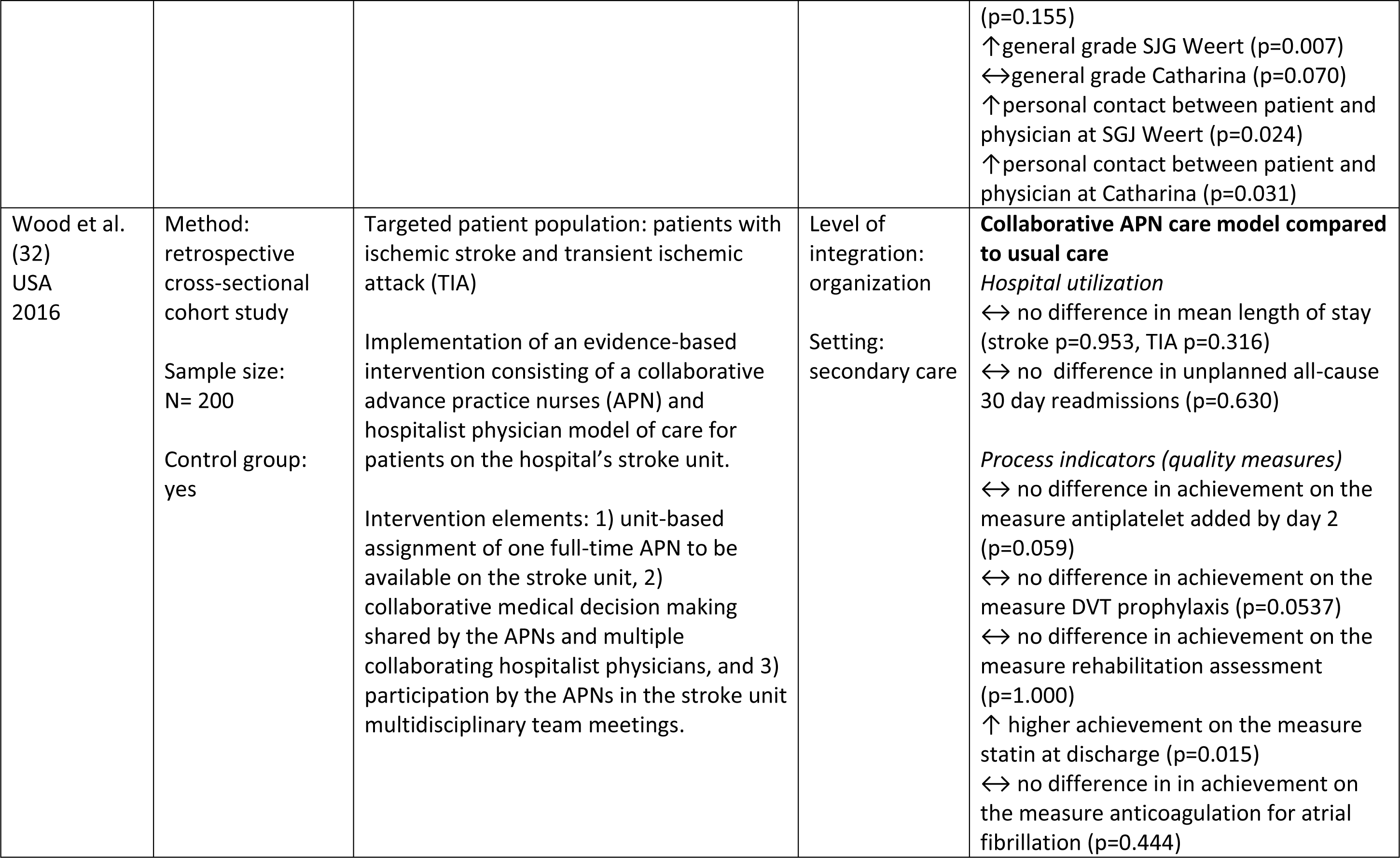
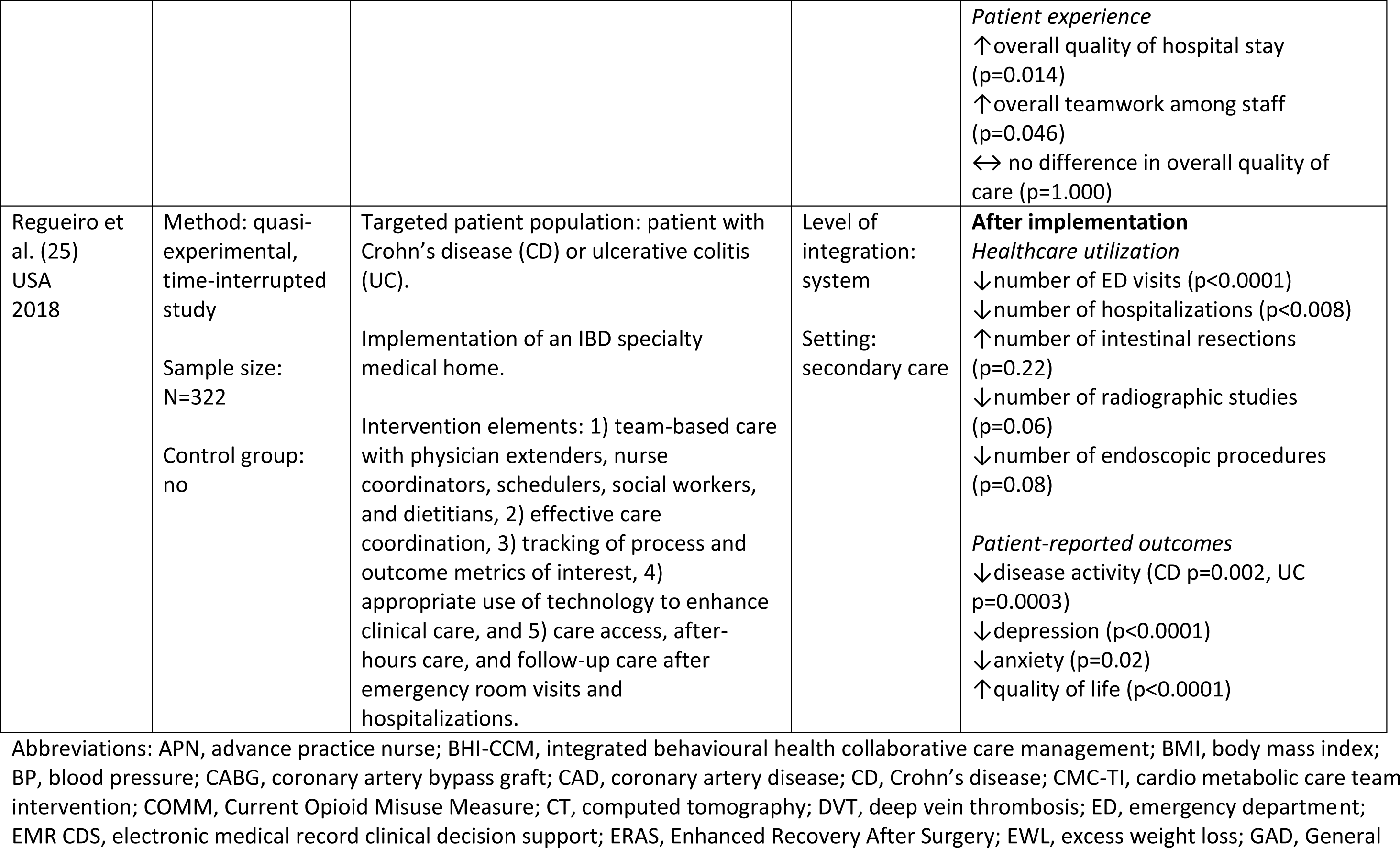
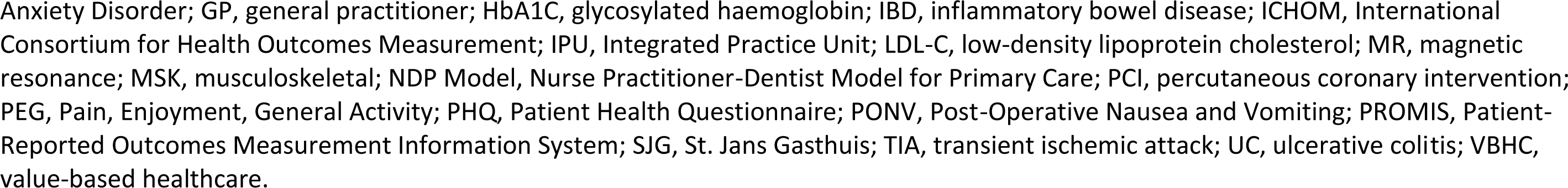
A description of the interventions and quantitative results of the value-based integrated care interventions.

#### 3.4.2 Effects of value-based integrated care – quantitative analysis

In eleven studies, a wide range of outcome measures was used to evaluate the effect of the VBIC intervention (Table 2). All articles analysed the effect of the VBIC intervention on multiple outcome measures. The outcome measures consisted of: 1) patient-reported outcomes (e.g. quality of life, disease activity), 2) clinical outcomes (e.g. HbA1c, weight, mortality), 3) healthcare utilization (e.g. emergency department (ED) visits, hospitalizations, patient encounters), 4) cost of care, 5) patient experiences (e.g. quality of care, satisfaction with care), and 6) process indicators (e.g. proportion of patients that received care according to protocol). Almost all articles described a positive effect of the VBIC intervention on at least one of these outcome measures. More specifically, multiple articles reported a positive impact of the intervention on the following: 1) HbA1c (26, 30, 31, 36), 2) number of ED visits (25, 28, 30), 3) number of hospitalizations (25, 28, 30), 4) number of endoscopies (25, 28), 5) number of radiographic studies (25, 28), and 6) quality of life (25, 34, 36). Across the different articles, no consensus was reached on the impact of the VBIC intervention on the cost of care (26, 28–30, 36), systolic blood pressure (30, 31), mortality (26, 27, 29), re-admission rate (26, 32) and quality of care (27, 32).

#### 3.4.3 Effects of value-based integrated care – qualitative analysis

One article(35) reports the results of a qualitative evaluation. This study by Nilsson et al. aimed to explore how participants experienced the implementation of VBHC at a Swedish University Hospital(35). A part of the intervention focused on increasing cooperation with other departments or care institutions within the care chain. This review focused on this part of the intervention, not the intervention as a whole. The participants noted that the increased cooperation across departments made it easier to obtain outcome measurements and to perform patient follow-ups. In addition, increased cooperation increased the participants understanding of different conditions treated at each department and of conditions for different patient populations. Furthermore, the intervention increased the awareness of cooperation between inpatient and outpatient care. The increase in cooperation contributed to increased accessibility for the patients to receive care at the right care level.(35)

### 3.5 Facilitators and barriers for the implementation of value-based integrated care

Almost all articles (n=19, 79%) described either a facilitator or barrier for the implementation of VBIC. The various facilitators and barriers for the implementation of VBIC were grouped into nine different categories: 1) information technology, 2) financing, 3) organizational culture and leadership, 4) workforce, 5) communication and coordination, 6) commitment, 7) clinical care, 8) education, and 9) quality improvement. Facilitators were most often mentioned in the categories of information technology, financing, and communication and coordination (Table 3). Specifically, the most frequently reported facilitators were supportive information technology (n=8), a new reimbursement or payment model (n=7) and leadership (n=4). Barriers were mentioned most often in the categories of information technology, financing and workforce. Commonly reported barriers were limited or insufficient information technology (n=8), current reimbursement or payment model (n=7), and the required cultural change (n=4).

**Table 3:**
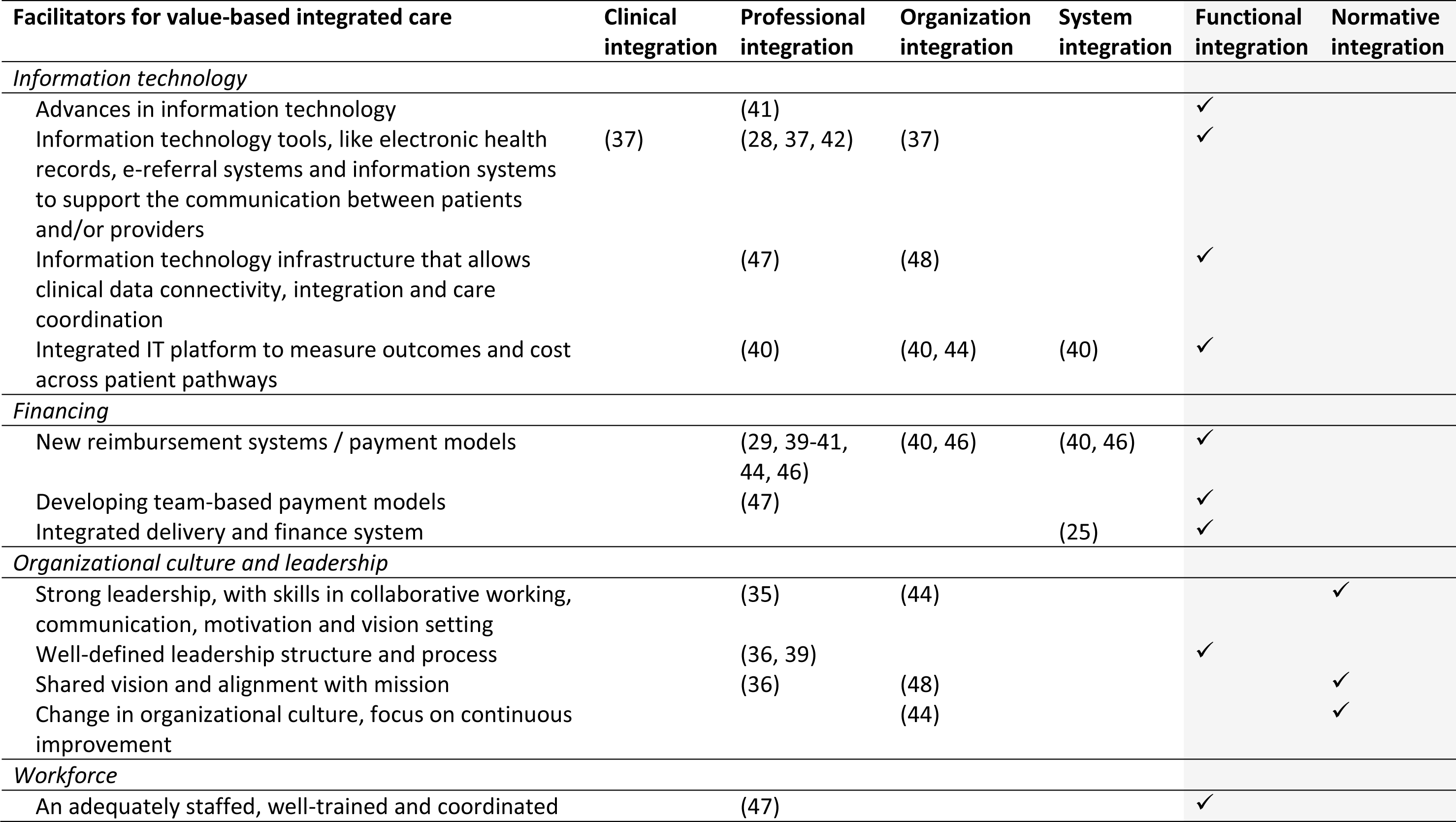
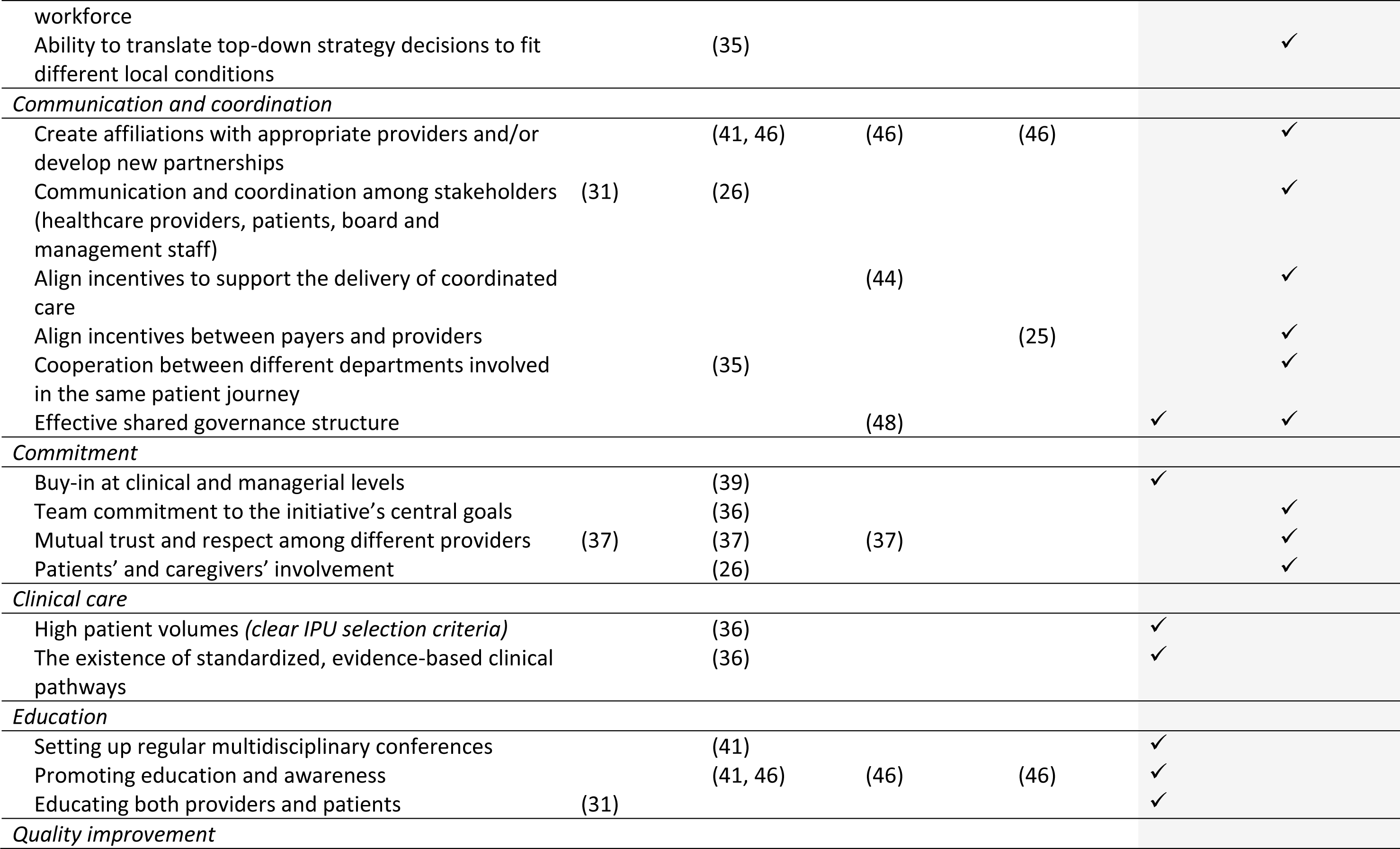
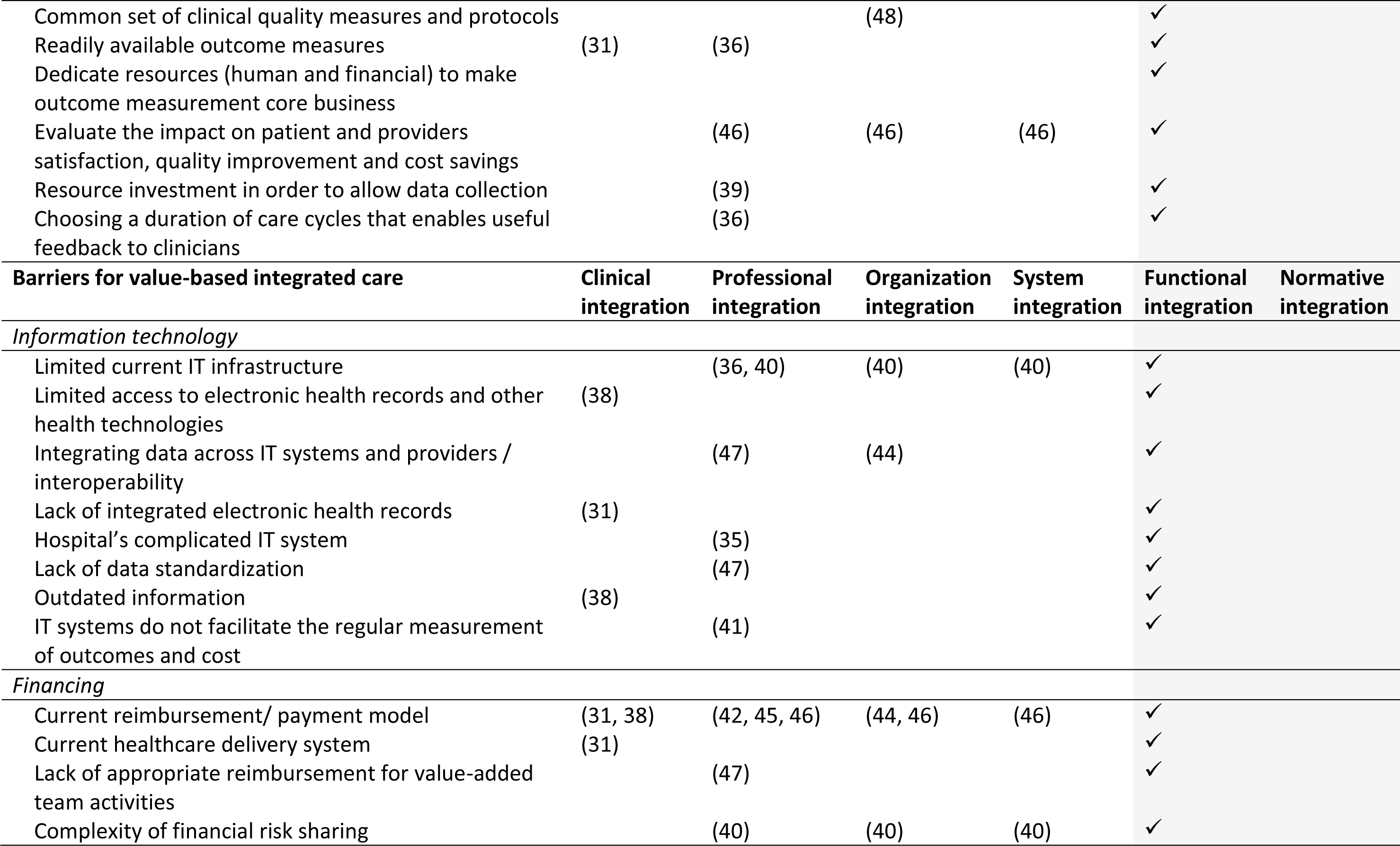
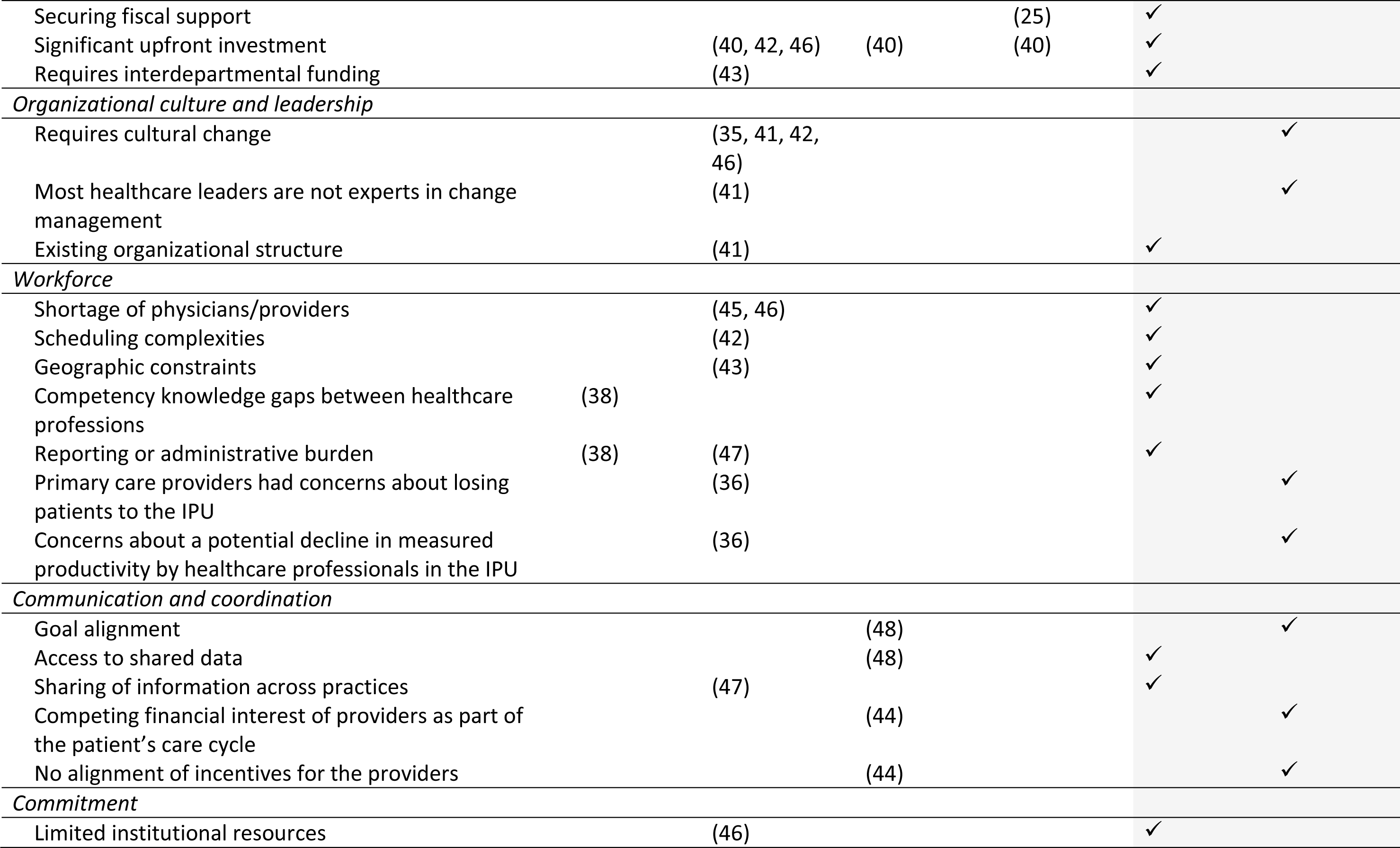
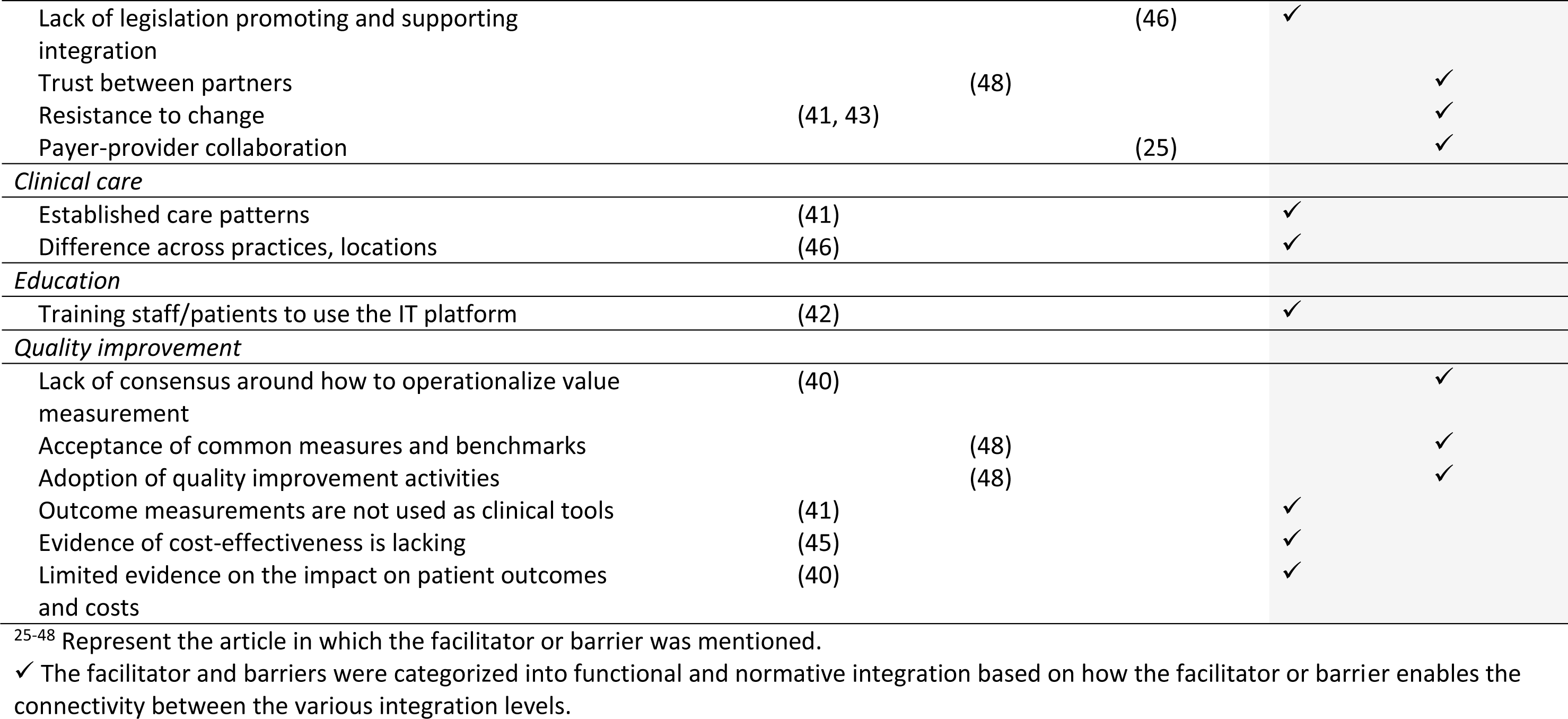
Facilitators and barriers for the implementation of value-based integrated care categorized according to the dimensions of the Rainbow Model of Integrated Care.

## 4. Discussion

The main objective of this study was to provide practical evidence-based recommendations for the implementation of integrated care within a VBHC context, in other words, value-based integrated care. To achieve this aim, we identified how VBIC is defined in current literature, summarized the results of evaluations of the effects of VBIC and summarized the literature regarding the facilitators and barriers for its implementation. Among the articles included in this systematic review, we found one definition for the concept of VBIC. This definition largely overlaps with the principles of VBHC. Two of the core elements of VBHC consist of maximizing patient value by measuring patient achieved outcomes in relation to the amount of money spent to achieve those outcomes, and organizing care around medical conditions and care cycles for a specific patient population. With the exception of patient experience with care, all elements mentioned within the definition of VBIC are also described within the VBHC framework. This confirms that VBIC fits within the larger context of VBHC; the additional focus on patient experiences can be understood from the context of integrated care, which aims to improve patient care and experiences through improved care coordination(5, 9).

How VBIC fits within the integrated care context is more difficult to distinguish. This is directly related to the complex nature of integrated care; possible similarities between the two concepts are dependent on the used definition of integrated care. However, regardless of the precise definition integrated care is, at its core, an approach to overcome care fragmentation(8). Providing accessible, comprehensive and coordinated care is therefore an important element of both integrated care and VBIC(8, 37).

Furthermore, we aimed to evaluate the effects of VBIC implementation. The results suggest that VBIC interventions may have a positive impact on clinical outcomes, patient-reported outcomes and healthcare utilization. Comparing the effects of VBIC with the effects of integrated care reveals many similarities. Previous systematic reviews on the effects of integrated care suggest that it might have a positive effect on hospital admissions(18), readmissions(18) and patient satisfaction(18, 19). This was confirmed in our systematic review regarding VBIC.

At last, this review assessed the facilitators and barriers for the implementation of VBIC. Overall, our findings highlight that healthcare organizations which aim to successfully implement VBIC must invest in a satisfactory IT infrastructure, support and facilitate the implementation of new reimbursement or payment models, remove barriers to cultural change and support strong leadership. A strong leader at the helm of the VBIC intervention can facilitate its implementation by creating an environment in which healthcare providers or organizations are stimulated to trust each other and work together to achieve their common goal. Supportive leadership can thereby also facilitate cultural change. Comparing the factors that influence the successful implementation of VBIC with the facilitating factors for the implementation of VBHC or integrated care reveals many similarities. A well-functioning IT infrastructure, financial support, leadership and cultural change are also frequently mentioned facilitators and barriers for the implementation of VBHC(49–52) or integrated care(16, 17, 53).

### 4.1 Strengths and limitations

This is the first systematic review about VBIC. Both empirical and non-empirical studies were included to obtain a broad overview of the current literature on VBIC. No distinction was made as to the type or level of VBIC, which enabled us to provide recommendations for the implementation of VBIC across the whole spectrum of integrated care.

However, several limitations should also be mentioned. Firstly, this review may not include all relevant articles on VBIC. Although the search strategy aimed to include all articles broadly related to VBIC, we might have missed articles that used different terms to describe VBIC. The absence of registered index terms (e.g. MeSH or Emtree) for VBHC, integrated care or VBIC also complicated the search. However, since the search strategy included all known terms, synonyms and spelling variations for VBHC, integrated care and VBIC, we expect the chance of missing a relevant article to be minimal.

Secondly, since the term VBIC is rarely used, and the definition was still unknown during article selection, the reviewers of this study decided that an article was considered to fit the criteria of VBIC if it described the implementation of integrated care within a VBHC context. This may have led to the inclusion of articles that did not strictly fit the definition of VBIC as provided by Valentijn et al.(37) In addition, the use of the self-determined VBIC criteria may have led to the inclusion of articles that were primarily about VBHC instead of VBIC. Nonetheless, we believe that we screened the articles very carefully and only included articles on VBIC.

Lastly, our findings on the effectiveness of VBIC interventions should be interpreted with caution. The generalizability is hindered by the limited number of studies that evaluated the effectiveness of a VBIC intervention and the different characteristics of the VBIC intervention. Each study implemented an intervention for a different target population, achieved a different level of integration and used different outcomes measures. Moreover, some articles used a large number of outcome measures and did not define one specific primary outcome measure to evaluate the effect of the intervention. Those articles often found at least one significant reduction or improvement on their outcome measures, which might have been a result of multiple testing.

### 4.2 Implications for research and practice

The reviews findings suggest that the concepts VBIC, VBHC and integrated care share a certain level of resemblance. Similarities can be found in the definitions, the effects and the facilitators and barriers for implementation. Resemblance is also inherent to the VBIC interventions; the interventions consist of multiple components related to both VBHC and integrated care. The resemblance between the three concepts, together with the multicomponent interventions, restrict our ability to assess causality between the separate components of the intervention and the results. The added value of VBIC above VBHC or integrated care, therefore, remains unclear. This raises the question if VBIC is substantially different from VBHC and integrated care, and if a separate definition for VBIC provides additional value. Evidently, it would be prudent to further investigate the resemblance and possible distinction between VBIC, VBHC and integrated care.

Moreover, clear guidelines should be developed to facilitate the implementation and evaluation of VBIC, VBHC or integrated care interventions. Those guidelines should include recommendations on research design and the selection of outcome measures. Based on our findings we recommend that further research should evaluate the effectiveness of an implementation using a randomized clinical trial, stepped-wedge or cohort design (i.e. a study design with a control group), to ensure that researchers are able to assess causality. In addition, we recommend that researchers evaluate the effects of one intervention at a time and critically assess the outcome measures needed to measure the effect, and define those upfront. All outcome measures must be relevant to the intervention and should be included if a change is expected to occur after implementation, to limit the burden of outcome collection and the possibility of multiple testing.

Furthermore, our findings provide an overview of all possible factors that influence the successful implementation of VBIC. Health organizations wishing to implement VBIC can use this overview to create the ideal environment for implementation and increase the chance of a successful implementation. Further research could be performed to identify the underlying mechanism of the influencing factors. Such research will increase the understanding of why a certain factor facilitates or hinders VBIC and provide more insight into the intricacies of VBIC implementation.

An increased understanding of the facilitating or hindering factors for VBIC is also necessary to enable organization to achieve sustainable healthcare services. To maximize patient value across the whole cycle of care, care needs to be integrated on organizational or preferably system level. Many VBIC initiatives, however, focus on achieving clinical or professional integration. These initiatives are often driven by a bottom-up approach, advocated by healthcare professionals with the aim to deliver more patient-centred care. The transformation to system level integration requires both a bottom-up and top-down approach; it requires collaboration between professionals, organizations, governments and healthcare insurers. This collaboration needs to be supported by (national) policies and regulations, and by functional and normative integration mechanisms such as a shared mission and adequate financial, management and information systems. Only by facilitating collaborations and removing the barriers for integration will healthcare organizations be able to achieve true VBIC on a system level.

## 5. Conclusions

This systematic review found that the concept of VBIC is not well defined in current literature. The effect of VBIC seems promising and comparable to integrated care or VBHC, but the exact interpretation of effect evaluations is challenged by the precedence of multicomponent interventions, multiple testing and generalizability issues. For successful implementation of integrated care within a VBHC context, it is imperative that healthcare organizations consider investing in appropriate IT infrastructure and the development and implementation of new reimbursement models.

## Declarations

### Authors’ contributions

ESvH led the development of the search strategy, screened all papers (first screener), led the analysis and wrote the first draft of the manuscript. LY was involved in screening of the papers (second screener), analysis, and reviewed the manuscript. NvL was involved in conceptualization of the study, development of the search strategy, screening of the papers (third screener), analysis, and reviewed the manuscript. HR was involved in conceptualization of the study, was involved in the analysis, and reviewed the manuscript. HFL led the conceptualization of the study, was involved in the analysis, and reviewed the manuscript. All authors approved the final version of the manuscript.

## Acknowledgements

We would like to thank our consortium members: I.L. Abma, C.T.B. Ahaus, R.J. Baatenburg de Jong, M.C. de Bruijne, M.C. Dorr, E.A.C. Dronkers, H.J. van Elten, L. Haverman, J.G.M. Jelsma, M. Leusder, M.M. van Muilekom, M.P.J. Offerman, T.S. Reindersma, K.S. van Hof, and P.J van der Wees for their contribution.

We also wish to thank W.M. Bramer from the Erasmus MC Medical Library for developing and updating the search strategies.

## Data availability statement

The authors confirm that the data supporting the findings of this study are available within the article and its supplementary materials.

