## Supplementary materials for "Value-Based Integrated Care: A Systematic Literature Review"

**1. Search string**

***Embase.com***

('integrated care'/de OR 'integrated health care system'/de OR (('integration'/de OR 'cooperation'/de) AND ('health care system'/de OR 'health care delivery'/de)) OR 'disease management'/de OR 'case management'/de OR 'interprofessional collaboration'/de OR 'multidisciplinary team'/de OR 'collaborative care team'/de OR 'accountable care organization'/de OR (((integrat* OR coordinat* OR collaborat* OR multidisciplin* OR multi-disciplin* OR accountab* OR comprehensive* OR  seamless* OR transmural* OR trans-mural*) NEAR/6 (care OR healthcare OR delivery OR service* OR team OR provision* OR case-model* OR patient*)) OR ((disease OR case) NEAR/3 management*) OR ((interprofession* OR inter-profession*) NEAR/3 (collaborat* OR coordinat*))):ab,ti) AND ('value based care'/de OR 'value based medicine'/de OR 'value-based insurance design'/de OR (value-based OR valuebased):ab,ti) NOT ([conference abstract]/lim  OR [letter]/lim OR [note]/lim OR [editorial]/lim)

***Medline ALL Ovid***

(Delivery of Health Care, Integrated/ OR Disease Management/ OR Case Management/ OR Accountable Care Organizations/ OR (((integrat* OR coordinat* OR collaborat* OR multidisciplin* OR multi-disciplin* OR accountab* OR comprehensive* OR  seamless* OR transmural* OR trans-mural*) ADJ6 (care OR healthcare OR delivery OR service* OR team OR provision* OR case-model* OR patient*)) OR ((disease OR case) ADJ3 management*) OR ((interprofession* OR inter-profession*) ADJ3 (collaborat* OR coordinat*))).ab,ti.) AND (Value-Based Health Insurance / OR Value-Based Purchasing / OR (value-based OR valuebased).ab,ti.)

***Web of Science Core Collection***

TS=(((((integrat* OR coordinat* OR collaborat* OR multidisciplin* OR multi-disciplin* OR accountab* OR comprehensive* OR  seamless* OR transmural* OR trans-mural*) NEAR/5 (care OR healthcare OR delivery OR service* OR team OR provision* OR case-model* OR patient*)) OR ((disease OR case) NEAR/2 management*) OR ((interprofession* OR inter-profession*) NEAR/2 (collaborat* OR coordinat*)))) AND ((value-based OR valuebased))) AND DT=(article)

***Cochrane CENTRAL register of Trials***

((((integrat* OR coordinat* OR collaborat* OR multidisciplin* OR multi-disciplin* OR accountab* OR comprehensive* OR  seamless* OR transmural* OR trans-mural*) NEAR/6 (care OR healthcare OR delivery OR service* OR team OR provision* OR case-model* OR patient*)) OR ((disease OR case) NEAR/3 management*) OR ((interprofession* OR inter-profession*) NEAR/3 (collaborat* OR coordinat*))):ab,ti) AND ((value-based OR valuebased):ab,ti)

**2. Results per database**

| **Database** | **Number of studies** | **Number of studies after deduplication** |
|---|---|---|
| Embase.com | 830 | 94 |
| Medline ALL Ovid | 957 | 948 |
| Web of Science Core Collection | 781 | 274 |
| Cochrane CENTRAL register of Trials | 42 | 12 |
| **Total** | **2610** | **1328** |
